# Predictors and barriers for the management of non-communicable diseases among older Syrian refugees amidst the COVID-19 pandemic in Lebanon: A cross-sectional analysis of a multi-wave survey

**DOI:** 10.1101/2022.04.12.22273786

**Authors:** Stephen J. McCall, Tanya El Khoury, Noura Salibi, Berthe Abi Zeid, Maria El Haddad, Marwan F. Alawieh, Sawsan Abdulrahim, Monique Chaaya, Hala Ghattas, Abla Sibai

**Affiliations:** Center for Research on Population and Health, Faculty of Health Sciences, American University of Beirut, Beirut, Lebanon; Department of Epidemiology and Population Health, Faculty of Health Sciences, American University of Beirut, Beirut, Lebanon; Norwegian Refugee Council (NRC), Beirut, Lebanon; Department of Health Promotion and Community Health, Faculty of Health Sciences, American University of Beirut, Beirut, Lebanon

**Keywords:** Non-Communicable Diseases, Chronic Diseases, Syrian Refugees, Older adults, Vulnerable populations, Lebanon

## Abstract

**Importance:** Older Syrian refugees have a high burden of non-communicable diseases and economic vulnerability.

**Objective:** This study aimed to develop and internally validate a predictive model of the inability to manage non-communicable diseases (NCDs) in older Syrian refugees, and to describe barriers to adherence to NCD medication.

**Design:** A nested cross-sectional study within a longitudinal study.

**Setting:** Lebanon.

**Population:** Syrian refugees aged 50 years or older residing in Lebanon who self-reported having hypertension, diabetes, history of cardiovascular disease (CVD) or chronic respiratory disease (CRD).

**Methods:** All households with refugees aged 50 years or older who received humanitarian assistance from a non-governmental organization were invited to participate in a study examining the impact of COVID-19 on older Syrian refugees. Data were collected through telephone surveys between September 2020 and January 2021. The study outcome was self-reported inability to manage hypertension, diabetes, CVD or CRD. Predictors of inability to manage any NCD were assessed using a logistic regression models. The model was internally validated using bootstrapping techniques, which gave an estimate of optimism. The optimism-adjusted discrimination and calibration of the model were presented using C-statistic and calibration slope (C-slope), respectively.

**Results:** Out of 3,222 older Syrian refugees, 1,893 reported having at least one NCD including 43% who had hypertension, 24% diabetes, 24% history of CVD, and 11% CRD. There were 387 (20%) participants who were unable to manage at least one of their NCDs. Predictors for inability to manage NCDs were age, non-receipt of cash assistance, household water insecurity, household food insecurity, and having multiple chronic diseases. The model’s adjusted C-statistic was 0.65 (95%CI:0.62-0.67) and C-slope was 0.88 (95%CI:0.73-1.03). The prevalence of non-adherence to medication was 9% and the main reasons were unaffordability of medication (41%) and the belief that they no longer required the medication after feeling better (22%).

**Conclusions:** This study identified that the predictors of inability to manage NCDs among older Syrian refugees in Lebanon are mainly related to financial barriers, which aids the targeting of assistance and interventions. Context-appropriate assistance is required to overcome financial barriers and enable equitable access to medication and healthcare.

**Key points:** *Question:* What are the predictors and barriers to managing NCDs as an older Syrian refugee in Lebanon?

*Findings:* This nested cross-sectional study assessed the predictors and barriers to managing any NCD, which included hypertension, diabetes, history of cardiovascular disease and chronic respiratory disease. Predictors included age, no cash assistance, household water insecurity, household food insecurity and having multiple chronic diseases. Primary reasons for not taking medications were unaffordability of the medication and belief medication was no longer required.

*Meaning:* Context-appropriate assistance is required to overcome financial barriers and enable equitable access to healthcare and medication required to manage NCDs.

## Introduction

Forcibly displaced populations have been recognized as vulnerable populations to the direct and indirect impacts of the COVID-19 pandemic.^1-3^ Furthermore, older refugees are particularly vulnerable as they have a higher prevalence of pre-existing chronic disease^4^ and have an increased risk of hospitalization and mortality due to COVID-19.^5^ Even prior to the COVID-19 pandemic, poor management of non-communicable diseases (NCDs) was a global public health concern.^6^ NCD self-management, including lifestyle changes and adherence to medication, has been shown to reduce premature mortality and morbidity.^7-9^ In refugee populations, however, NCD management is problematic due to the competing demands of basic needs such as access to adequate food, shelter, protection and water. Furthermore, NCD self-management in refugee populations is exacerbated by poor access to health services, language barriers between the patient and the healthcare provider, being food insecure, and having limited health literacy.^10^

In Lebanon, large populations of Syrian refugees are situated in the Bekaa valley and North Lebanon and reside in informal tented settlements or residential areas.^11^ These are areas where the Lebanese health system was already stretched to meet the needs of the host population, and since 2011 have faced unprecedented challenges in meeting the needs of Syrian refugees.^12^ Refugees have access to subsidized healthcare services and medication mainly through the United Nations High Commissioner for Refugees (UNHCR); however, despite these subsidies they are often unable to cover the full cost of needed medication.^13^ The COVID-19 pandemic and the recent major economic crisis in Lebanon have impacted access to healthcare and supply of essential medicines^14^, which is likely to further exacerbate refugees’ abililty to manage chronic conditions.

Studies on the management of NCDs among refugees in Lebanon and the Middle East, a region that hosts one of the largest refugee populations, are scarce.^15^ Understanding the barriers and predictors of self-reported management of NCDs among older refugees, a population at high-risk, is important to allow resource allocation and contextualized humanitarian assistance to prevent premature mortality and morbidity. The present study aimed to elucidate the predictors of inability to manage NCDs in older Syrian refugees and describe barriers to accessing healthcare and managing these chronic conditions.

## Methods

### Study design, sample and study population

This was a cross-sectional study nested within a multi-wave longitudinal study, which aimed to examine the vulnerabilities of older Syrian refugees residing in Lebanon during the COVID-19 pandemic.^16^ The study included Syrian refugees aged 50 years and older, who were identified from a full listing of beneficiary households of a non-governmental humanitarian organization [Norwegian Refugee Council].

Within the beneficiary sampling frame, all households who had used services offered by the humanitarian organization between 2017 and 2020 and included an adult aged 50 years or older were contacted and were included in the study. If there were multiple adults aged 50 years or older within a household, one person was randomly selected and was assessed for capacity to consent, invited to participate, and provided oral consent to enter the study. The same respondent was approached to complete a telephone survey across different time points. The data for the present analysis were extracted from Waves 1 (September 2020-December 2020) and Waves 2 (October 2020-January 2021). This study was reported according to the TRIPOD reporting guideline for prediction modelling.^17^

Out of 17,384 households initially contacted at wave 1, 4,010 eligible beneficiary Syrian refugees aged 50 years and over were invited to participate; of those, 3,322 beneficiaries consented and participated in both waves 1 and 2 (Figure 1), 1893 respondents reported having at least one NCD and and 387 reported being unable to manage at least one NCD.

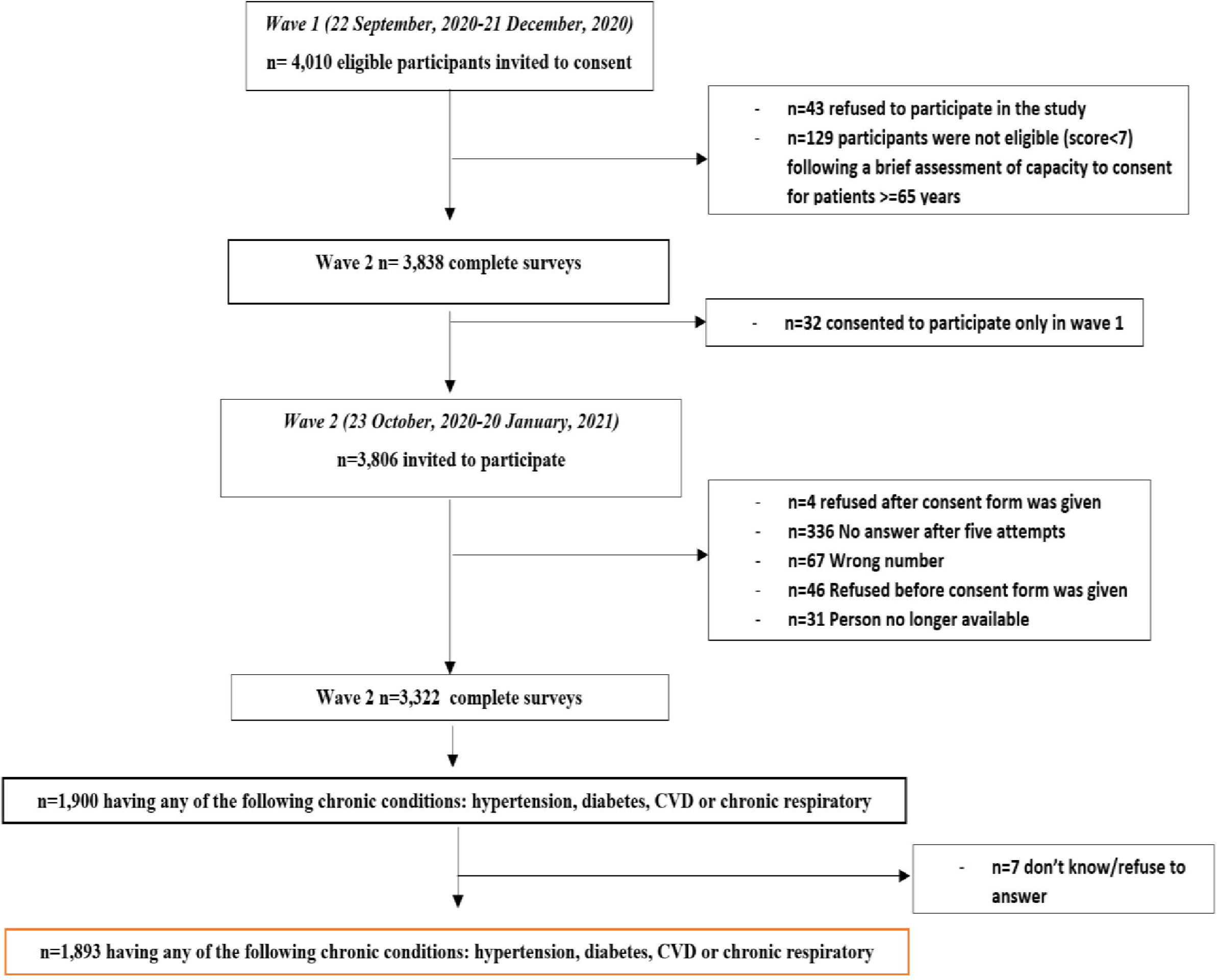

### Data sources

The questionnaire for each wave was developed using a combination of sources including validated questionnaire modules, contextually specific questions, and community-identified priorities. The survey was co-created by academics, humanitarian actors, local government officials, and focal points from the refugee communities. Modules varied between the waves; the demographic module was in wave 1, and the NCD and access to healthcare module was included in wave 2. The questionnaire was piloted internally with data collectors and local community focal points to ensure face validity. Trained data collectors administered the surveys in Arabic and entered data into structured electronic data collection forms hosted on Kobo toolbox. Data entry checks and monitoring were performed for quality assurance.

### Outcome Measures

The outcome variable was self-reported inability to manage any NCD. This included the following conditions: hypertension, diabetes, cardiovascular disease or chronic respiratory disease. For each condition, participants were asked the following question “Are you able to manage your [Insert condition]?”.

### Candidate predictors

Using the literature, 16 potential predictors were identified for inability to manage NCDs and were included for model development. These included: age (continuous); sex (male/female); residence (outside/inside tented settlements); education (preparatory and elementary/never attended); smoking status (smoker/ex-smoker/never-smoker); number of chronic conditions (one/two/three or more); hypertension (yes/no); diabetes (yes/no); chronic respiratory disease (yes/no); cardiovascular disease (yes/no); living arrangement (alone/with someone); food insecurity measured using the Food Insecurity Experience Scale ^18^(severe food insecurity (raw score 7-8)/mild-moderate food insecurity (raw score 4-6)/ food secure raw score 0-3)); employment status (yes/no); water insecurity measured using the short-form Household Water Insecurity Scale^19^ (yes/no); receipt of cash or voucher assistance (yes/no) and family debts (yes/no).

### Missing data

The largest amount of missing data in any variable was 1.8%, and these were assumed to be missing at random, so we used complete case analysis.^20^

### Statistical Analysis

Absolute frequencies and proportions were presented alongside odds ratios and their 95% confidence intervals using unadjusted logistic regression models, which examined the odds of inability to manage any NCD for each candidate predictor.

All variables were categorical other than age, which had a linear relationship with inability to manage NCD. All candidate predictors of inability to manage NCD were entered into multivariable logistic regression models and removed using a stepwise backwards method using a P<0.157 ^21^. This has been used as a proxy for the Akaike Information Criterion (AIC), where predictors are removed to obtain the lowest AIC. Multicollinearity was assessed using correlation matrices and variance inflation factor; a variance inflation factor > 5 indicated collinearity. Number of NCDs and each separate chronic condition were modelled in two separate models. Smoking status was removed from the final model to improve model fit and prevent overfitting.^22^

The final model’s performance in terms of discrimination was assessed using the C-statistic, which ranges from 0.5 – 1.0 (this is also known as the area under the receiver operating characteristic curve (AUC-ROC)), where a value of 1.0 represents perfect discriminative ability between those with and without the outcome, and 0.5 denotes a discriminative ability equal to chance. We also assessed the calibration of the final model, which describes the agreement between observed outcomes and predictive probabilities.^23^ This was assessed using calibration plots, which categorizes patients into ten groups according to predictive probabilities, where the mean predicted risk within each of these groups is plotted against the mean observed proportion of events.^23,24^ If there is a perfect calibration, the graph will show a diagonal line where there is a slope of 1 and intercept of 0. A slope <1 suggests overfitting in the model, this is where respondents with high risk of the outcome have overestimated risk predictions while those with low risk of the outcome have underestimated risk predictions. In addition, we assessed whether there was an overall difference between the observed number of events and the average predictive risk using the calibration-in-the-large.

The final model’s selection of predictors, discrimination and calibration estimates were internally validated using bootstrap methods; where 500 bootstrap samples with replacement were used to validate the model selection process and generate an estimate of optimism, optimism adjusted estimates of C-statistic and optimism-adjusted calibration plot were generated. ^20,23,25,26^ Bootstrap shrinkage was applied to the final apparent model and the modified beta coefficients and odds ratios were presented. All analyses were conducted using Stata/SE 17.

### Ethical approval

This study was approved by the American University of Beirut Social and Behavioral Sciences Institutional Review Board [Reference: SBS-2020-0329]. Refugees are considered vulnerable populations, and extensive efforts were made to ensure autonomy was respected throughout the data collection.

## Results

### Characteristics of the population

Out of a total of 1,893 participants, 387 noted that they were unable to manage their condition through any means (lifestyle or medication) (Table 1). Among these participants, 174 were unable to manage their NCD through medication. The study sample median age was 59 (IQR: 54-65), and 1089 (58%) were women. Hypertension topped the listed of reported NCDs (74%), this was followed by cardiovascular disease (42%), diabetes (41%), and chronic respiratory disease (19%).

**Table 1.**
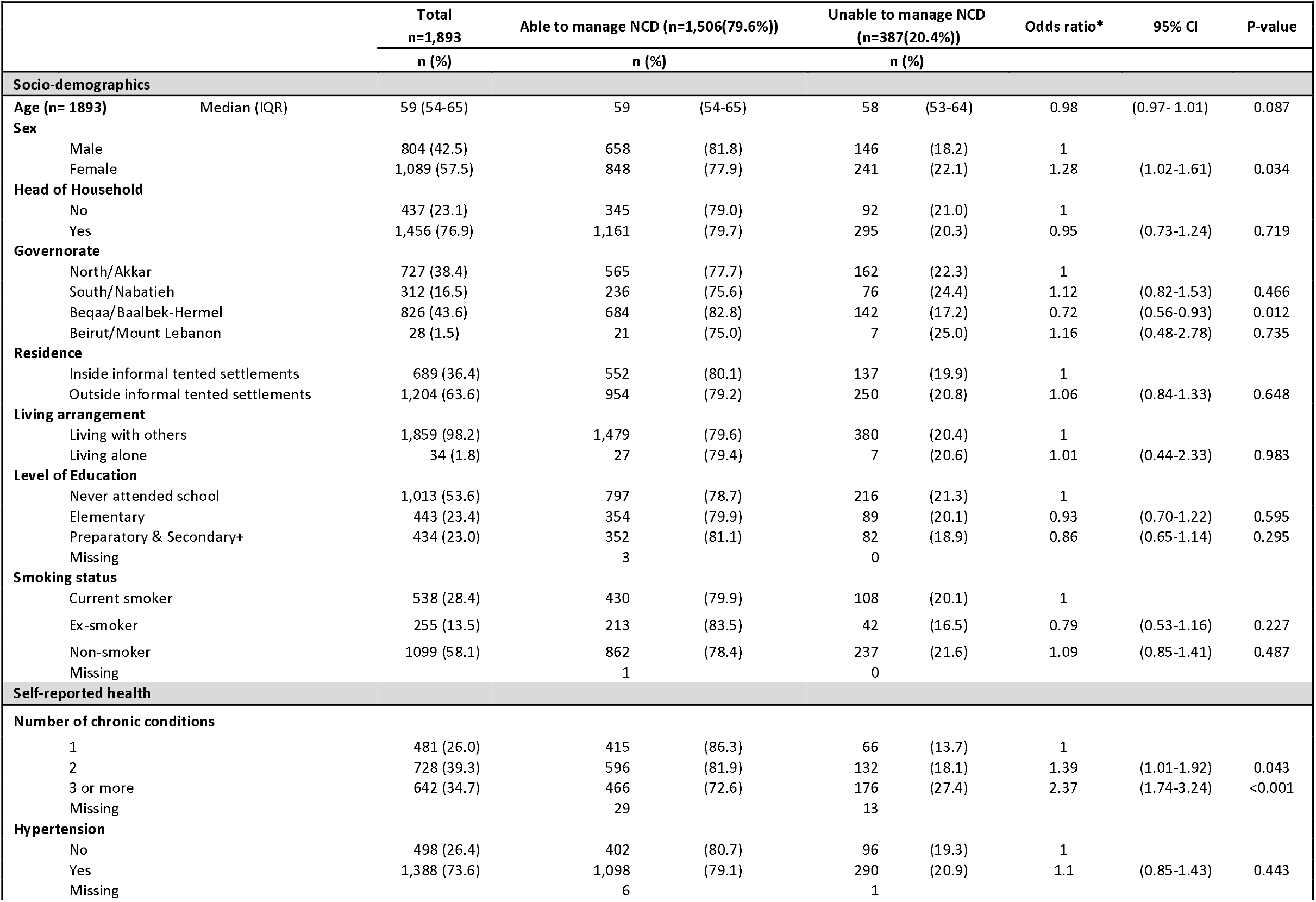
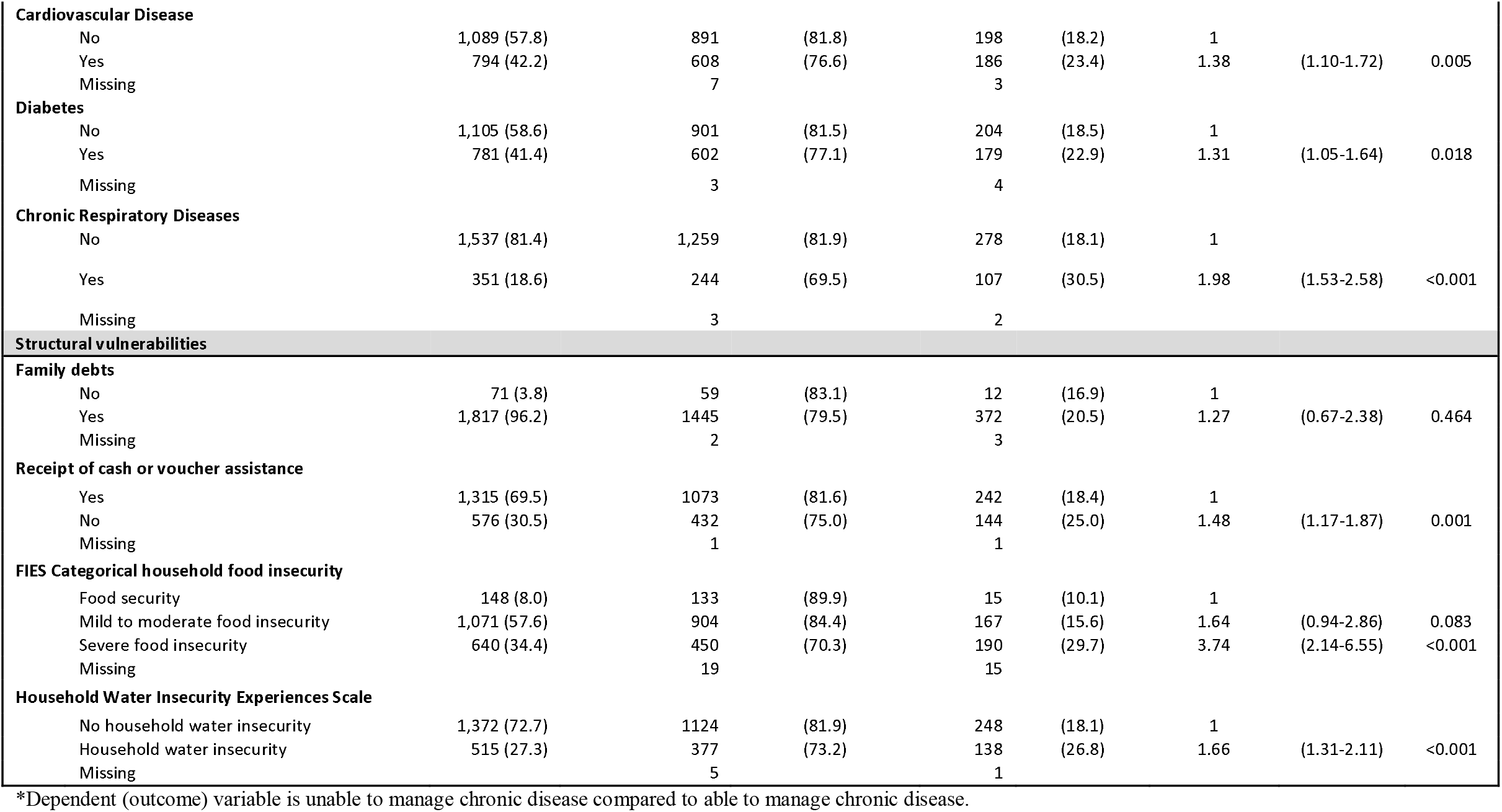
Characteristics and vulnerability markers of older Syrian refugees by the ability to manage their non-communicable disease

|  |  | Total<br>n=1,893 | Able to manage NCD (n=1,506(79.6%)) |  | Unable to manage NCD<br>(n=387(20.4%)) |  | Odds ratio* | 95% CI | P-value |
| --- | --- | --- | --- | --- | --- | --- | --- | --- | --- |
|  |  | n (%) | n (%) |  | n (%) |  |  |  |  |
| Socio-demographics |  |  |  |  |  |  |  |  |  |
| Age (n= 1893) | Median (IQR) | 59 (54-65) | 59 | (54-65) | 58 | (53-64) | 0.98 | (0.97- 1.01) | 0.087 |
| Sex |  |  |  |  |  |  |  |  |  |
|  | Male | 804 (42.5) | 658 | (81.8) | 146 | (18.2) | 1 |  |  |
|  | Female | 1,089 (57.5) | 848 | (77.9) | 241 | (22.1) | 1.28 | (1.02-1.61) | 0.034 |
| Head of Household |  |  |  |  |  |  |  |  |  |
|  | No | 437 (23.1) | 345 | (79.0) | 92 | (21.0) | 1 |  |  |
|  | Yes | 1,456 (76.9) | 1,161 | (79.7) | 295 | (20.3) | 0.95 | (0.73-1.24) | 0.719 |
| Governorate |  |  |  |  |  |  |  |  |  |
|  | North/Akkar | 727 (38.4) | 565 | (77.7) | 162 | (22.3) | 1 |  |  |
|  | South/Nabatieh | 312 (16.5) | 236 | (75.6) | 76 | (24.4) | 1.12 | (0.82-1.53) | 0.466 |
|  | Beqaa/Baalbek-Hermel | 826 (43.6) | 684 | (82.8) | 142 | (17.2) | 0.72 | (0.56-0.93) | 0.012 |
|  | Beirut/Mount Lebanon | 28 (1.5) | 21 | (75.0) | 7 | (25.0) | 1.16 | (0.48-2.78) | 0.735 |
| Residence |  |  |  |  |  |  |  |  |  |
|  | Inside informal tented settlements | 689 (36.4) | 552 | (80.1) | 137 | (19.9) | 1 |  |  |
|  | Outside informal tented settlements | 1,204 (63.6) | 954 | (79.2) | 250 | (20.8) | 1.06 | (0.84-1.33) | 0.648 |
| Living arrangement |  |  |  |  |  |  |  |  |  |
|  | Living with others | 1,859 (98.2) | 1,479 | (79.6) | 380 | (20.4) | 1 |  |  |
|  | Living alone | 34 (1.8) | 27 | (79.4) | 7 | (20.6) | 1.01 | (0.44-2.33) | 0.983 |
| Level of Education |  |  |  |  |  |  |  |  |  |
|  | Never attended school | 1,013 (53.6) | 797 | (78.7) | 216 | (21.3) | 1 |  |  |
|  | Elementary | 443 (23.4) | 354 | (79.9) | 89 | (20.1) | 0.93 | (0.70-1.22) | 0.595 |
|  | Preparatory & Secondary+ | 434 (23.0) | 352 | (81.1) | 82 | (18.9) | 0.86 | (0.65-1.14) | 0.295 |
|  | Missing |  | 3 |  | 0 |  |  |  |  |
| Smoking status |  |  |  |  |  |  |  |  |  |
|  | Current smoker | 538 (28.4) | 430 | (79.9) | 108 | (20.1) | 1 |  |  |
|  | Ex-smoker | 255 (13.5) | 213 | (83.5) | 42 | (16.5) | 0.79 | (0.53-1.16) | 0.227 |
|  | Non-smoker | 1099 (58.1) | 862 | (78.4) | 237 | (21.6) | 1.09 | (0.85-1.41) | 0.487 |
|  | Missing |  | 1 |  | 0 |  |  |  |  |
| Self-reported health |  |  |  |  |  |  |  |  |  |
| Number of chronic conditions |  |  |  |  |  |  |  |  |  |
|  | 1 | 481 (26.0) | 415 | (86.3) | 66 | (13.7) | 1 |  |  |
|  | 2 | 728 (39.3) | 596 | (81.9) | 132 | (18.1) | 1.39 | (1.01-1.92) | 0.043 |
|  | 3 or more | 642 (34.7) | 466 | (72.6) | 176 | (27.4) | 2.37 | (1.74-3.24) | <0.001 |
|  | Missing |  | 29 |  | 13 |  |  |  |  |
| Hypertension |  |  |  |  |  |  |  |  |  |
|  | No | 498 (26.4) | 402 | (80.7) | 96 | (19.3) | 1 |  |  |
|  | Yes | 1,388 (73.6) | 1,098 | (79.1) | 290 | (20.9) | 1.1 | (0.85-1.43) | 0.443 |
|  | Missing |  | 6 |  | 1 |  |  |  |  |

|  |  |  |  |  |  |  |  |  |
| --- | --- | --- | --- | --- | --- | --- | --- | --- |
| **Cardiovascular Disease** |  |  |  |  |  |  |  |  |
| No | 1,089 (57.8) | 891 | (81.8) | 198 | (18.2) | 1 |  |  |
| Yes | 794 (42.2) | 608 | (76.6) | 186 | (23.4) | 1.38 | (1.10-1.72) | 0.005 |
| Missing |  | 7 |  | 3 |  |  |  |  |
| **Diabetes** |  |  |  |  |  |  |  |  |
| No | 1,105 (58.6) | 901 | (81.5) | 204 | (18.5) | 1 |  |  |
| Yes | 781 (41.4) | 602 | (77.1) | 179 | (22.9) | 1.31 | (1.05-1.64) | 0.018 |
| Missing |  | 3 |  | 4 |  |  |  |  |
| **Chronic Respiratory Diseases** |  |  |  |  |  |  |  |  |
| No | 1,537 (81.4) | 1,259 | (81.9) | 278 | (18.1) | 1 |  |  |
| Yes | 351 (18.6) | 244 | (69.5) | 107 | (30.5) | 1.98 | (1.53-2.58) | <0.001 |
| Missing |  | 3 |  | 2 |  |  |  |  |
| **Structural vulnerabilities** |  |  |  |  |  |  |  |  |
| **Family debts** |  |  |  |  |  |  |  |  |
| No | 71 (3.8) | 59 | (83.1) | 12 | (16.9) | 1 |  |  |
| Yes | 1,817 (96.2) | 1445 | (79.5) | 372 | (20.5) | 1.27 | (0.67-2.38) | 0.464 |
| Missing |  | 2 |  | 3 |  |  |  |  |
| **Receipt of cash or voucher assistance** |  |  |  |  |  |  |  |  |
| Yes | 1,315 (69.5) | 1073 | (81.6) | 242 | (18.4) | 1 |  |  |
| No | 576 (30.5) | 432 | (75.0) | 144 | (25.0) | 1.48 | (1.17-1.87) | 0.001 |
| Missing |  | 1 |  | 1 |  |  |  |  |
| **FIES Categorical household food insecurity** |  |  |  |  |  |  |  |  |
| Food security | 148 (8.0) | 133 | (89.9) | 15 | (10.1) | 1 |  |  |
| Mild to moderate food insecurity | 1,071 (57.6) | 904 | (84.4) | 167 | (15.6) | 1.64 | (0.94-2.86) | 0.083 |
| Severe food insecurity | 640 (34.4) | 450 | (70.3) | 190 | (29.7) | 3.74 | (2.14-6.55) | <0.001 |
| Missing |  | 19 |  | 15 |  |  |  |  |
| **Household Water Insecurity Experiences Scale** |  |  |  |  |  |  |  |  |
| No household water insecurity | 1,372 (72.7) | 1124 | (81.9) | 248 | (18.1) | 1 |  |  |
| Household water insecurity | 515 (27.3) | 377 | (73.2) | 138 | (26.8) | 1.66 | (1.31-2.11) | <0.001 |
| Missing |  | 5 |  | 1 |  |  |  |  |

\*Dependent (outcome) variable is unable to manage chronic disease compared to able to manage chronic disease.

Among 174 participants who were unable to manage their NCD through medication, the primary reasons included unaffordability of the medication (40.8%), feeling better and not needing the medication (22.4%) and medication not always available (14.4%). Unaffordability of medication remained the primary reason for hypertension, diabetes and cardiovascular disease when examining disease-specific reasons for non-adherence to medication (Figure 2a). In addition, there were 295 participants who were unable to manage their NCDs and unable to access primary health care. The primary reason for being unable to access primary care in this group was the cost of the doctor’s visits, medication or tests (80.7%) (Figure 2b).

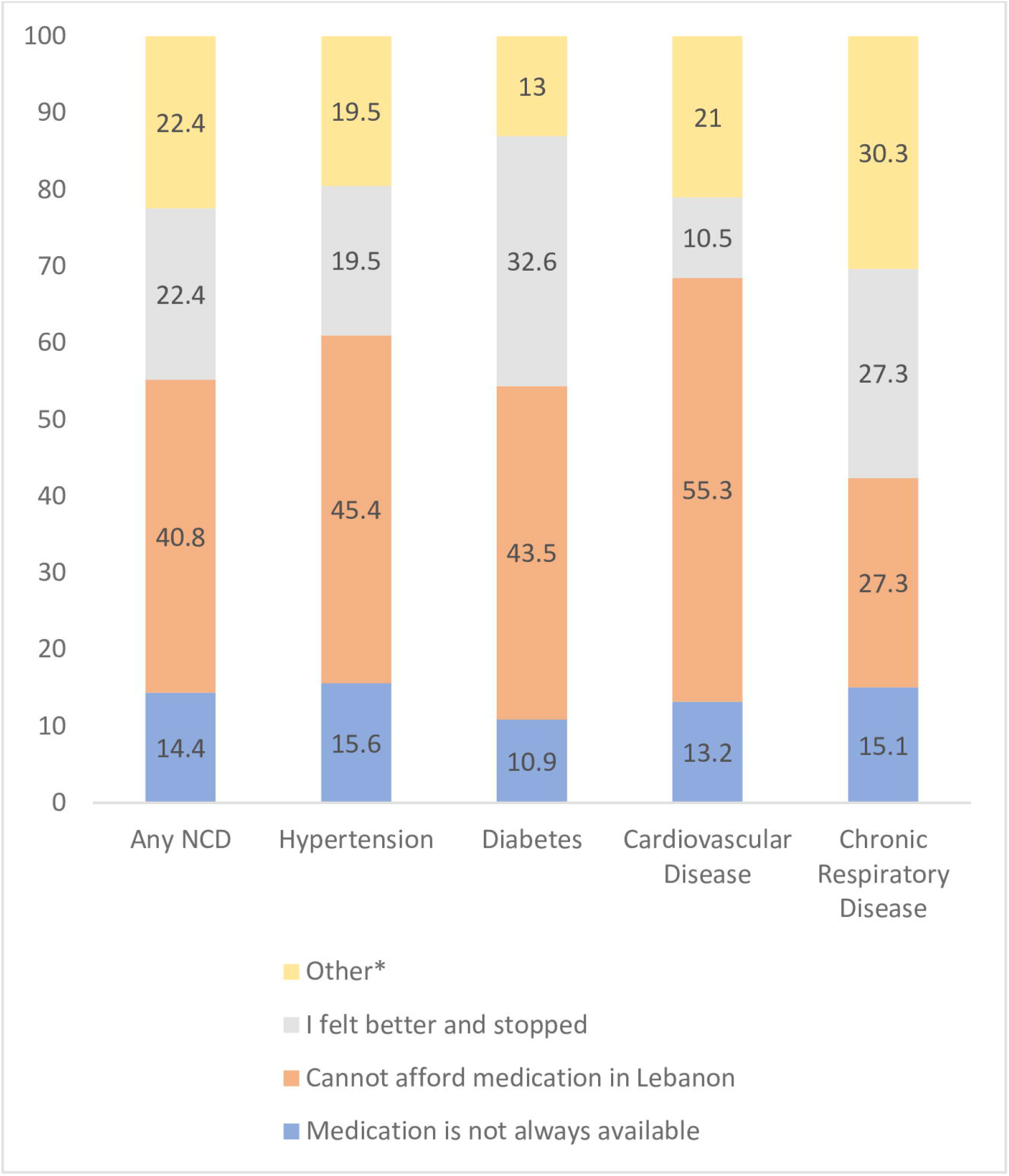
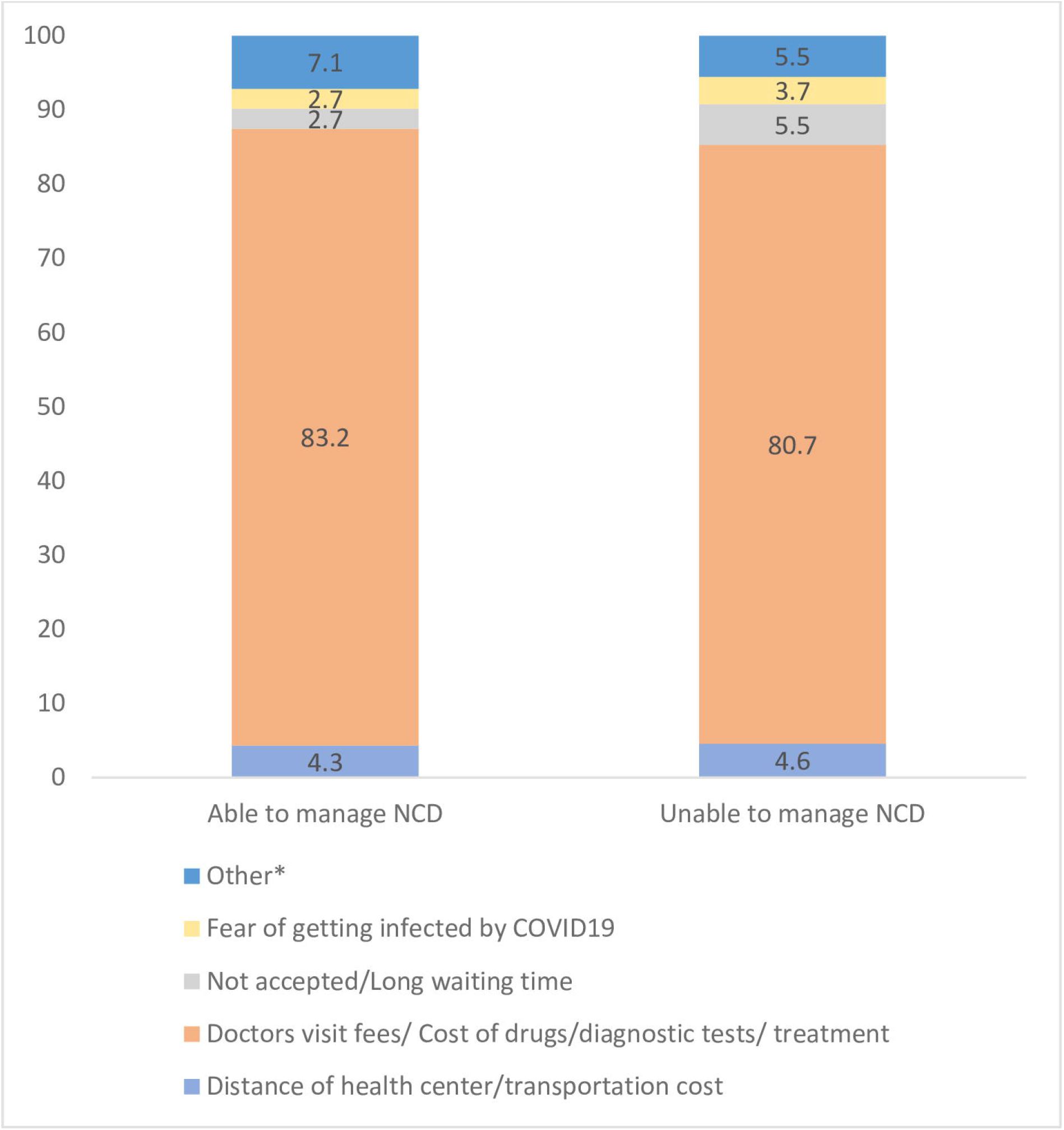

### Unadjusted analysis

Unadjusted odds ratios of inability to manage NCD and 95% confidence interval of each potential predictor are reported in Table 1. This study identified different characteristics among older refugees that had increased odds of being unable to manage at least one NCD. These included being female compared to male (OR:1.28 (95% CI: 1.02-1.61)), having multiple chronic conditions (OR:1.39 (95% CI: 1.01-1.92)) compared to having none, having a history of cardiovascular disease (OR:1.38 (95% CI: 1.10-1.72)), diabetes (OR:1.31 (95% CI: 1.05-1.64)) or chronic respiratory disease (OR:1.98 (95% CI: 1.53-2.58) compared to not having an NCD, not having received cash (OR:1.48 (95% CI: 1.17-1.87)) compared to receipt of assistance, and having experienced severe food insecurity compared to food security (OR:3.74 (95% CI: 2.14-6.55)) and household water insecurity (OR:1.66 (95%CI: 1.31-2.11)) compared to being water secure.

### Predictors and model performance

The final model retained eight predictors of inability to manage NCD, which included age, self-reported hypertension, diabetes, cardiovascular disease and chronic respiratory disease (the more NCDs an individual possesses the higher the predicted risk of inability to manage them), severe household food insecurity, no receipt of cash assistance, and household water insecurity (Table 2). The final model had a moderate discriminative ability with an optimized adjusted C-statistic of 0.65 (95% CI: 0.62 – 0.68) (Table 2 and Figure 3a). The calibration plot without correction for optimism is shown in Figure 3b and the optimism adjusted calibration plot is shown in Figure 3a which depicts the expected performance of the final model in future samples.^23,27^ The calibration slope after adjustment for optimism was 0.88 (95% CI: 0.73 - 1.03) (Table 2 and Figure 3a). The odds ratios and coefficients of the final model have been adjusted for overfitting and are presented in Table 2.

**Table 2.** Multivariable model for predicting inability to manage any NCD

|  | Apparent model |  |  |  |  | Model adjusted by bootstrap shrinkage |  |  |  |  |
| --- | --- | --- | --- | --- | --- | --- | --- | --- | --- | --- |
|  | Parameter estimate | (95%CI) | Odds Ratio | 95% CI | P-value | Parameter estimate | (95% CI) | Odds Ratio | 95% CI | P-value |
| **Age** | -0.01 | (-0.03 - 0.00) | 0.99 | (0.97-1.00) | 0.14 | -0.01 | (-0.02 - 0.00) | 0.99 | (0.98-1.00) | 0.142 |
| **Self-reported medical history** |  |  |  |  |  |  |  |  |  |  |
| Hypertension | 0.35 | (0.06 - 0.64) | 1.42 | (1.07 - 1.89) | 0.02 | 0.31 | (0.06 - 0.56) | 1.36 | (1.06-1.75) | 0.016 |
| Diabetes | 0.35 | (0.11 - 0.60) | 1.42 | (1.11 - 1.81) | <0.001 | 0.31 | (0.09 -0.52) | 1.36 | (1.10-1.68) | 0.005 |
| Cardiovascular disease | 0.22 | (-0.02 - 0.46) | 1.24 | (0.98 - 1.58) | 0.08 | 0.19 | (-0.02 - 0.40) | 1.21 | (0.98-1.50) | 0.075 |
| Chronic respiratory disease | 0.8 | (0.50 - 1.09) | 2.22 | (1.66 - 2.96) | <0.001 | 0.70 | (0.44 - 0.95) | 2.01 | (1.56-2.59) | <0.001 |
| **FIES Categorical household food insecurity** |  |  |  |  |  |  |  |  |  |  |
| Food secure | Ref |  |  |  |  | Ref |  |  |  |  |
| Mild to moderate food insecurity | 0.5 | (-0.08 - 1.08) | 1.64 | (0.92 - 2.94) | 0.09 | 0.44 | (-0.07 - 0.94) | 1.55 | (0.93-2.57) | 0.094 |
| Severe food insecurity | 1.21 | (0.62 - 1.80) | 3.37 | (1.87 - 6.07) | <0.001 | 1.06 | (0.55 - 1.58) | 2.90 | (1.73-4.85) | <0.001 |
| **Household Water Insecurity Experiences Scale** |  |  |  |  |  |  |  |  |  |  |
| No household water insecurity | Ref |  |  |  |  | Ref |  |  |  |  |
| Household water insecurity | 0.2 | (-0.06 - 0.46) | 1.22 | (0.94 - 1.59) | 0.14 | 0.17 | (-0.06 - 0.40) | 1.19 | (0.95-1.50) | 0.137 |
| **Receipt of cash or voucher assistance** |  |  |  |  |  |  |  |  |  |  |
| Yes | Ref |  |  |  |  | Ref |  |  |  |  |
| No | 0.3 | (0.05 - 0.55) | 1.35 | (1.05 - 1.73) | 0.02 | 0.26 | (0.04 - 0.48) | 1.30 | (1.04-1.62) | 0.02 |
| **Intercept** | -2.48 | (-3.63 - -1.33) | 0.08 | (0.03 - 0.26) | <0.001 | -2.33 | (-2.45 - -2.22) | 0.10 | (0.09-0.11) | <0.001 |
| **Calibration and discrimination of models** |  |  |  |  |  |  |  |  |  |  |
|  | 95% CI |  |  |  |  | 95% CI |  |  |  |  |
| **C-statistic /Area under the curve** | 0.67 | (0.638-0.701) |  |  |  | 0.651 | (0.621 - 0.677) |  |  |  |
| **C-slope** | 1 | (0.805-1.195) |  |  |  | 0.876 | (0.728 - 1.033) |  |  |  |
| **Calibration in the large** | 0 | (-0.119 - 1.119) |  |  |  | 0.005 | (-0.111 0.128) |  |  |  |

\*Reference category for each self-reported medical condition is “no”.

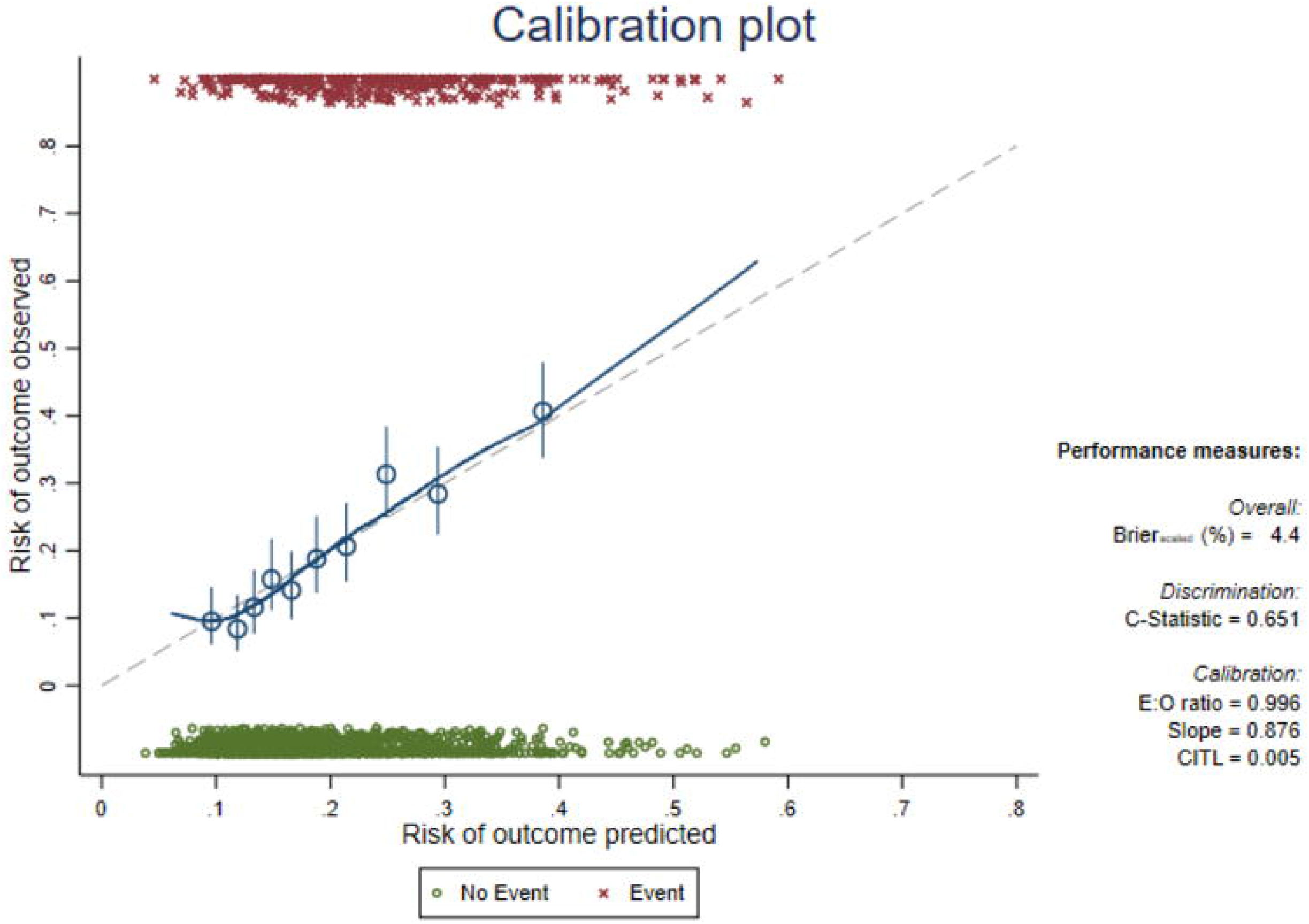
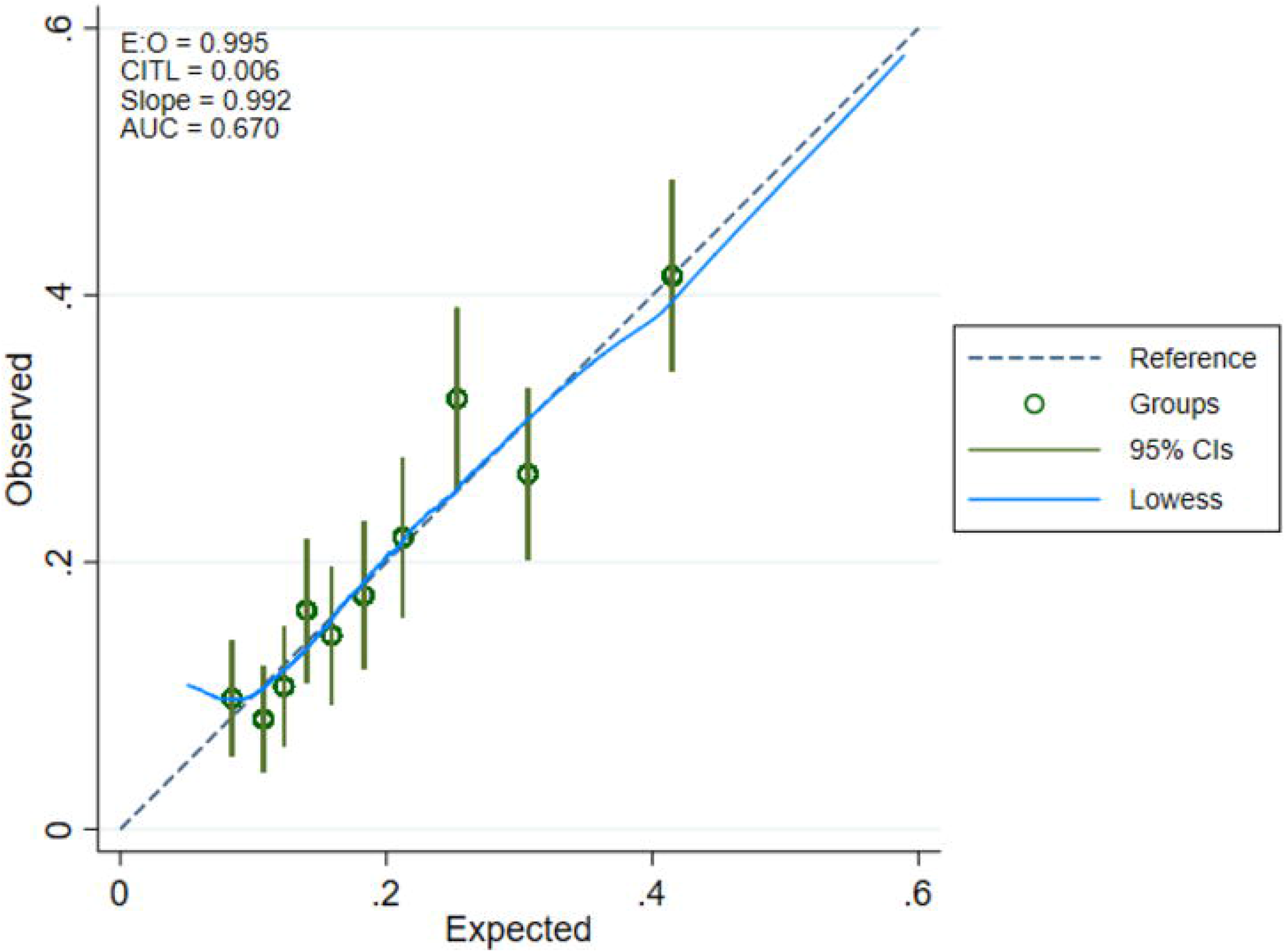

All the included predictors had the expected direction coefficient with the outcome. In particular, receiving no cash assistance, having household water insecurity or severe food insecurity had positive coefficients, and hence increased the likelihood of older Syrian refugees being unable to manage their NCDs. Age had a small negative coefficient with inability to manage NCDs.

To illustrate, the predicted risk of inability to manage NCDs for a Syrian refugee aged 60 years and over, with all four NCDs, severe household food insecurity, no receipt of cash assistance and household water insecurity was 51% (using the optimized-adjusted coefficients). Whilst the predicted risk of inability to manage NCDs for a Syrian refugee aged 60 years old with all four NCDs, who has received cash assistance and had no food insecurity and no household water insecurity was 19% (using the optimized-adjusted coefficients). For a Syrian refugee aged 60 years or old who had hypertension and diabetes without other NCDs the predicted risk was 9%.

In a sensitivity analysis, total number of chronic diseases was modelled as a categorical variable rather than each individual NCD. The predictors of this alternative model are: the number of chronic diseases, food insecurity and non-receipt of cash assistance. However, the discrimination (C-Statistics: 0.63 (95% CI: 0.60-0.66)) and calibration (C-Slope 0.86 (95%CI: 0.71-1.06)) of the optimized-adjusted model were poorer than the final model (Supplementary table 1).

## Discussion

This study has identified predictors of inability to manage NCDs among older Syrian refugees in Lebanon during the COVID-19 pandemic. Younger age, not receiving cash assistance, household water insecurity, severe household food insecurity and co-morbidity were predictors of inability to manage NCDs. One of the key barriers for older Syrian refugees to adhere to medication was cost of the medication and beliefs that they did not require the medication anymore after feeling better. In addition, for older Syrians who were unable to access primary healthcare, the cost of the visit, tests or medications was the main reason.

### Results in context

Our study showed that Syrian refugees who did not receive cash assistance had a higher likelihood of being unable to manage their NCDs. This finding concurs with a study conducted in Lebanon among Syrian refugees showing that receiving multipurpose cash assistance (MPC) lead to an increased access to primary health care for various illnesses, including chronic diseases.^28^ The majority of Syrian refugees rely on cash assistance to cover their basic needs such as food, rent, water and healthcare.^29^ Hence, one possible explanation is that without cash assistance resources are limited to the acute and immediate livelihood needs rather than managing a chronic disease. According to UNHCR, medications for chronic diseases are provided with a small handling fee to Syrian refugees in Lebanon. However, those with conditions requiring long term, high cost treatments, mainly related to chronic diseases, are not covered, which is a main gap to be addressed in the humanitarian response.^12^ Furthermore, our results showed that affordability was the primary reason for not taking chronic disease medication or attending primary healthcare. Similarly, studies performed in Lebanon and Jordan among Syrian refugees showed that the inability to afford cost of the treatment and medications primarily prevent seeking health care for chronic disease.^13,30^ It remains crucial for humanitarian agencies to remove the financial barriers for older Syrian populations in accessing required healthcare and essential chronic disease medication.

Water insecurity and severe food insecurity among Syrian refugees are markers of severe economic vulnerability and low socioeconomic status.^31-33^ As a result, households that are unable to meet their basic needs for food and water are unlikely to be able to use a proportion of their household income to pay for subsidized NCD medication. Furthermore, food insecurity has been shown to reduce overall dietary quality and diversity.^34-36^ Thus, food insecurity prevents refugees from modifying and improving their diet quality to manage their chronic disease.

Multimorbidity is common in older adults and it has implications on the self-management of these conditions.^37^ Our results are consistent with previous research, which showed that older adults with multimorbidity had reduced adherence to the management of their NCDs.^38^ Polypharmacy and complex dosing regimens are common in older age,^39^ these reduce compliance^38^ as each medication may have its own special instructions to follow,^40^ which can be increasingly difficult in older age.

Although there was a statistical association between younger age and self-management of NCDs, the magnitude of the estimand was small. Previous studies have shown that younger age was associated with lower medication adherence for depression^41^ and heart failure^42^. While other studies have suggested that there is no association between age and chronic medication adherence.

There is growing recognition that management of NCDs among refugee populations represents a challenge for humanitarian agencies, as these are costly to manage with limited resources available for healthcare ^13^. Further advocacy is required to donor organizations to prioritize NCDs in this population so that the health and wellbeing of refugee older population can be maximized. With the recent and unfortunate war in Ukraine and worsening of humanitarian catastrophe, there have been growing calls for immediate and rapid access to medicines for individuals with NCDs.^43^ In this context, reinforcing self-management protocols and health literacy, alleviating food insecurity and enabling dietary diversity are required to prevent secondary complications of unmanaged NCDs. Moreover, further studies targeting NCD interventions among this vulnerable population in the Middle East are crucial to reducing the disease burden espcailly in view of the very high prevalence of these conditions.

### Strengths and Limitations

The present study has enhanced our understanding of the predictors of inability to manage NCDs among older Syrian refugees in Lebanon and fills a major gap in the international literature. Furthermore, this study is one of the largest on older Syrian refugees in the published literature with a high response rate of >85% among those who were eligible.^11^ The study was limited as the predictive model had a moderate discriminative ability, which may be explained by missing predictors, such as perception of medication regime as being complicated, not knowing the purpose of the medication, accessibility issues, side effects due to medication, lack of healthcare support, lack of trust in doctors, stressful living conditions and time since diagnosis^15,30,44,45^. Furthermore, the calibration of the model showed overfitting and may not perform well in future samples, hence, future studies should aim to be larger if they wish to develop a predictive model. Another limitation was that data were self-reported to data collectors so misclassification in the data is possible; however, we tried to limit this through data quality and consistency checks.

### Conclusions

This study highlights that inability to manage NCDs was mainly related to financial barriers. The predictors from this study will allow healthcare professionals and humanitarian organizations to identify older refugees who are at a greater risk of being unable to manage their NCDs. These vulnerable groups should have the necessary assistance and support to allow an improvement in medication adherence and equitable access to healthcare. Furthermore, investment in NCD healthcare services in primary care will be beneficial to the prevention of premature mortality and morbidity from NCDs in Lebanon and elsewhere.

## Data Availability

All data produced in the present study are available upon reasonable request to the authors

## Acknowledgements

We thank Lara Abou Ammar, Nadine Rashidi, Zeina El Khoury, Zeinab Ramadan and Stephanie Bassil for their assistance in the study.

## References

1. World Health Organization (WHO). ApartTogether survey: preliminary overview of refugees and migrants self-reported impact of COVID-19. https://apps.who.int/iris/handle/10665/337931. Published 2020. Accessed 18 February, 2022.

2. Bukuluki P, Mwenyango H, Katongole SP, Sidhva D, Palattiyil G. The socio-economic and psychosocial impact of Covid-19 pandemic on urban refugees in Uganda. Social Sciences & Humanities Open. 2020;2(1):100045.

3. Fouad FM, McCall SJ, Ayoub H, Abu-Raddad LJ, Mumtaz GR. Vulnerability of Syrian refugees in Lebanon to COVID-19: quantitative insights. Conflict and Health. 2021;15(1):1–6.

4. Rehr M, Shoaib M, Ellithy S, et al. Prevalence of non-communicable diseases and access to care among non-camp Syrian refugees in northern Jordan. Conflict and Health. 2018;12(1):1–14.

5. Wang B, Li R, Lu Z, Huang Y. Does comorbidity increase the risk of patients with COVID-19: evidence from meta-analysis. Aging (Albany NY). 2020;12(7):6049.

6. WHO. Adherence to long term therapies: Evidence for action. http://apps.who.int/iris/bitstream/handle/10665/42682/9241545992.pdf;jsessionid=6B7994A65CA1BBDE3B6CB3ACDEDA4F49?sequence=1. Published 2003. Accessed 18 February, 2021.

7. Unal B, Critchley JA, Capewell S. Explaining the decline in coronary heart disease mortality in England and Wales between 1981 and 2000. Circulation. 2004;109(9):1101–1107.

8. Capewell S, Beaglehole R, Seddon M, McMurray J. Explanation for the decline in coronary heart disease mortality rates in Auckland, New Zealand, between 1982 and 1993. Circulation. 2000;102(13):1511–1516.

9. Nelson MR, Reid CM, Ryan P, Willson K, Yelland L. Self℧reported adherence with medication and cardiovascular disease outcomes in the second Australian National Blood Pressure Study (ANBP2). Medical Journal of Australia. 2006;185(9):487–489.

10. UNHCR. Promoting treatment adherence for refugees and persons of concern in healthcare settings. https://www.unhcr.org/5cd962bc7.pdf. Published 2019. Accessed 1 March, 2021.

11. DeJong J, Ghattas H, Bashour H, Mourtada R, Akik C, Reese-Masterson A. Reproductive, maternal, neonatal and child health in conflict: a case study on Syria using Countdown indicators. BMJ Global Health. 2017;2(3):e000302.

12. UNHCR. Health programme. https://www.unhcr.org/lb/wp-content/uploads/sites/16/2019/04/Health-Factsheet.pdf. Published 2019. Accessed 16 February 2022.

13. Doocy S, Lyles E, Akhu-Zaheya L, Burton A, Burnham G. Health service access and utilization among Syrian refugees in Jordan. International Journal for Equity in Health. 2016;15(1):1–15.

14. WHO. COVID-19 Significantly Impacts Health Services for Noncommunicable Diseases: World Health Organisation. https://www.who.int/news-room/detail/01-06-2020-covid-19-significantly-impacts-health-services-for-noncommunicable-diseases Published 2020. Accessed February 28, 2022.

15. Mohamad M, Moussally K, Lakis C, et al. Self-reported medication adherence among patients with diabetes or hypertension, Médecins Sans Frontières Shatila refugee camp, Beirut, Lebanon: A mixed-methods study. PloS ONE. 2021;16(5):e0251316.

16. Abdulrahim A, Ghattas H, McCall S. Changing vulnerabilities and COVID-19 adherence: older refugees in Lebanon. https://www.elrha.org/project/covid-19-adherence-older-refugees-lebanon/. Published 2020. Accessed 23 February 2022.

17. Moons KG, Altman DG, Reitsma JB, et al. Transparent Reporting of a multivariable prediction model for Individual Prognosis or Diagnosis (TRIPOD): explanation and elaboration. Annals of Internal Medicine. 2015;162(1):W1–W73.

18. Cafiero C, Viviani S, Nord M. Food security measurement in a global context: The food insecurity experience scale. Measurement. 2018;116:146–152.

19. Young SL, Miller JD, Frongillo EA, Boateng GO, Jamaluddine Z, Neilands TB. Validity of a four-item household water insecurity experiences scale for assessing water issues related to health and well-being. The American journal of tropical medicine and hygiene. 2021;104(1):391.

20. Harrell FE. Multivariable modeling strategies. In: Regression modeling strategies. Springer; 2015:63–102.

21. Sauerbrei W. The use of resampling methods to simplify regression models in medical statistics. Journal of the Royal Statistical Society: Series C (Applied Statistics). 1999;48(3):313–329.

22. Van Calster B, McLernon DJ, Van Smeden M, Wynants L, Steyerberg EW. Calibration: the Achilles heel of predictive analytics. BMC medicine. 2019;17(1):1–7.

23. Steyerberg EW. Overfitting and optimism in prediction models. In: Clinical Prediction Models. Springer; 2019:95–112.

24. Leijdekkers J, Eijkemans M, Van Tilborg T, et al. Predicting the cumulative chance of live birth over multiple complete cycles of in vitro fertilization: an external validation study. Human Reproduction. 2018;33(9):1684–1695.

25. Harrell FE, Lee KL, Mark DB. Multivariable prognostic models: issues in developing models, evaluating assumptions and adequacy, and measuring and reducing errors. Statistics in Medicine. 1996;15(4):361–387.

26. Fernandez-Felix B, García-Esquinas E, Muriel A, Royuela A, Zamora J. Bootstrap internal validation command for predictive logistic regression models. The Stata Journal. 2021;21(2):498–509.

27. Moons K, Donders ART, Steyerberg E, Harrell F. Penalized maximum likelihood estimation to directly adjust diagnostic and prognostic prediction models for overoptimism: a clinical example. Journal of Clinical Epidemiology. 2004;57(12):1262–1270.

28. Chaaban J, Salti N, Ghattas H, et al. Multi-purpose cash assistance in Lebanon: Impact evaluation on the well-being of Syrian refugees. American University of Beirut Press https://www.aubedulb/fafs/agri/aedrg/Documents/AUB%20Impact%20Study_Final_printpdf. 2020.

29. Ratnayake R, Rawashdeh F, AbuAlRub R, et al. Access to care and prevalence of hypertension and diabetes among Syrian refugees in northern Jordan. JAMA network open. 2020;3(10):e2021678–e2021678.

30. Lyles E, Burnham G, Chlela L, Spiegel P, Morlock L, Doocy S. Health service utilization and adherence to medication for hypertension and diabetes among Syrian refugees and affected host communities in Lebanon. Journal of Diabetes & Metabolic Disorders. 2020;19(2):1245–1259.

31. Workman CL, Brewis A, Wutich A, Young S, Stoler J, Kearns J. Understanding biopsychosocial health outcomes of syndemic water and food insecurity: applications for global health. The American Journal of Tropical Medicine and Hygiene. 2021;104(1):8.

32. Jamaluddine Z, Sahyoun NR, Choufani J, Sassine AJ, Ghattas H. Child-reported food insecurity is negatively associated with household food security, socioeconomic status, diet diversity, and school performance among children attending UN Relief and Works Agency for Palestine Refugees schools in Lebanon. The Journal of Nutrition. 2019;149(12):2228–2235.

33. Omidvar N, Ghazi-Tabatabie M, Sadeghi R, Mohammadi F, Abbasi-Shavazi MJ. Food insecurity and its sociodemographic correlates among Afghan immigrants in Iran. Journal of Health, Population, and Nutrition. 2013;31(3):356.

34. Leung CW, Epel ES, Ritchie LD, Crawford PB, Laraia BA. Food insecurity is inversely associated with diet quality of lower-income adults. Journal of the Academy of Nutrition and Dietetics. 2014;114(12):1943-1953. e1942.

35. Ghattas H, Sassine AJ, Seyfert K, Nord M, Sahyoun NR. Food insecurity among Iraqi refugees living in Lebanon, 10 years after the invasion of Iraq: data from a household survey. British Journal of Nutrition. 2014;112(1):70–79.

36. Seligman HK, Laraia BA, Kushel MB. Food insecurity is associated with chronic disease among low-income NHANES participants. The Journal of Nutrition. 2010;140(2):304–310.

37. Barnett K, Mercer SW, Norbury M, Watt G, Wyke S, Guthrie B. Epidemiology of multimorbidity and implications for health care, research, and medical education: a cross-sectional study. The Lancet. 2012;380(9836):37–43.

38. Kardas P, Lewek P, Matyjaszczyk M. Determinants of patient adherence: a review of systematic reviews. Frontiers in Pharmacology. 2013;4:91.

39. Linjakumpu T, Hartikainen S, Klaukka T, Veijola J, Kivelä S-L, Isoaho R. Use of medications and polypharmacy are increasing among the elderly. Journal of Clinical Epidemiology. 2002;55(8):809–817.

40. Ingersoll KS, Cohen J. The impact of medication regimen factors on adherence to chronic treatment: a review of literature. Journal of Behavioral Medicine. 2008;31(3):213–224.

41. Bambauer KZ, Soumerai SB, Adams AS, Zhang F, Ross-Degnan D. Provider and patient characteristics associated with antidepressant nonadherence: the impact of provider specialty. The Journal of Clinical Psychiatry. 2007;68(6):15841.

42. Bagchi AD, Esposito D, Kim M, Verdier J, Bencio D. Utilization of, and adherence to, drug therapy among medicaid beneficiaries with congestive heart failure. Clinical Therapeutics. 2007;29(8):1771–1783.

43. Ioffe Y, Abubakar I, Issa R, Spiegel P, Kumar BN. Meeting the health challenges of displaced populations from Ukraine. Lancet 2022.

44. Chew SM, Lee JH, Lim SF, Liew MJ, Xu Y, Towle RM. Prevalence and predictors of medication non℧adherence among older community℧dwelling people with chronic disease in Singapore. Journal of Advanced Nursing. 2021;77(10):4069–4080.

45. Nair KV, Belletti DA, Doyle JJ, et al. Understanding barriers to medication adherence in the hypertensive population by evaluating responses to a telephone survey. Patient Preference and Adherence. 2011;5:195.

