## Supplementary table 1 for "Predictors and barriers for the management of non-communicable diseases among older Syrian refugees amidst the COVID-19 pandemic in Lebanon: A cross-sectional analysis of a multi-wave survey"

**SUPPLEMENTARY MATERIALS**

Supplementary Table 1. Sensitivity analysis using the number of chronic conditions for the predictive model estimates and statistics

|  | **Apparent model** | | | | |  | **Bootstrap shrinkage adjusted model** | | | | |
| --- | --- | --- | --- | --- | --- | --- | --- | --- | --- | --- | --- |
|  | **Parameter estimate** | **(95%CI)** | **Odds Ratio** | **95% CI** | **P-value** |  | **Parameter estimate** | **(95% CI)** | **Odds Ratio** | **95% CI** | **P-value** |
| **Age** | -0.01 | [-0.03-0.00] | 0.99 | [0.97-1.00] | 0.09 |  | -0.01 | (-0.03-0.001) | 0.99 | (0.97-1.00) | 0.09 |
| **Number of chronic conditions** |  |  |  |  |  |  |  |  |  |  |  |
| 1 |  |  | 1 |  |  |  |  |  |  |  |  |
| 2 | 0.3 | [-0.04-0.63] | 1.34 | [0.96-1.88] | 0.08 |  | 0.26 | (-0.03-0.54) | 1.29 | (0.97-1.72) | 0.082 |
| 3 or more | 0.77 | [0.44-1.10] | 2.16 | [1.55-3.00] | <0.001 |  | 0.66 | (0.38-0.95) | 1.94 | (1.46-2.58) | <0.001 |
| **FIES Categorical household food insecurity** |  |  |  |  |  |  |  |  |  |  |  |
| Food secure |  |  | 1 |  |  |  |  |  |  |  |  |
| Mild to moderate | 0.52 | [-0.06-1.10] | 1.68 | [0.94-3.01] | 0.08 |  | 0.45 | (-0.05-0.95) | 1.57 | (0.95-2.59) | 0.078 |
| Severe food insecurity | 1.19 | [0.61-1.78] | 3.3 | [1.84-5.92] | <0.001 |  | 1.03 | (0.53-1.53) | 2.80 | (1.69-4.64) | <0.001 |
| **Receipt of cash or voucher assistance** |  |  |  |  |  |  |  |  |  |  |  |
| Yes |  |  | 1 |  |  |  |  |  |  |  |  |
| No | 0.28 | [0.03-0.53] | 1.32 | [1.03-1.70] | 0.03 |  | 0.24 | (0.03-0.46) | 1.27 | (1.03-1.58) | 0.027 |
| ***Intercept*** | -1.85 | [-2.96 - -0.74] | 0.16 | [0.05-0.48] | <0.001 |  | -1.79 | (-1.90 - 1.67) | 0.17 | (0.15-0.19) | <0.001 |
| ***Calibration and discrimination of models*** | | | | | | | | | | | |
|  |  | **95% CI** |  |  |  |  |  | **95% CI** |  |  |  |
| ***C-statistic /Area under the curve*** | 0.6523 | (0.621-0.683) |  |  |  |  | 0.633 | (0.602-0.662) |  |  |  |
| ***C-slope*** | 0 | (-0.118-0.118) |  |  |  |  | 0.863 | (0.710-1.064) |  |  |  |
| **Calibration in the large** | 1 | (0.778-1.222) |  |  |  |  | 0.006 | (-0.106-1.26) |  |  |  |
